# Motivation and Cognitive Abilities as Mediators between Polygenic Scores and Psychopathology in Children

**DOI:** 10.1101/2020.06.08.20123877

**Authors:** Narun Pat, Lucy Riglin, Richard Anney, Yue Wang, Deanna M. Barch, Anita Thapar, Argyris Stringaris

**Affiliations:** University of Otago, New Zealand, 9016; Cardiff University, UK, CF10 3AT; Washington University, USA, 63130; National Institute of Mental Health, USA, 20892

**Keywords:** Polygenic Score, Transdiagnostic Psychopathology, MDD, ADHD, Adolescent Brain Cognitive Development

## Abstract

**Objective:** Fundamental questions in biological psychiatry concern the mechanisms that mediate between genetic liability and psychiatric symptoms. Genetic liability for many common psychiatric disorders often confers transdiagnostic risk to develop a wide variety of psychopathological symptoms through yet unknown pathways. We examine the psychological and cognitive pathways that might mediate the relationship between genetic liability (indexed by polygenic scores; PS) and broad psychopathology (indexed by *p* factor and its underlying dimensions).

**Method:** We first identified which of the common psychiatric PSs (major depressive disorder [MDD], attention-deficit/hyperactivity disorder [ADHD], anxiety, bipolar, schizophrenia, autism) were associated with *p* factor. We then focused on three pathways: punishment sensitivity (reflected by behavioral inhibition system; BIS), reward sensitivity (reflected by behavioral activation system; BAS) and cognitive abilities (reflected by *g* factor based on 10 neurocognitive tasks). We applied structural equation modeling on the Adolescent Brain Cognitive Development (ABCD) dataset (n=4,814; 2,263 female children; 9-10 years old).

**Results:** MDD and ADHD PSs were associated with *p* factor. The association between MDD PS and psychopathology was partially mediated by punishment sensitivity and cognitive abilities: proportion mediated= 22.35%. Conversely, the influence of ADHD PS on psychopathology was partially mediated by reward sensitivity and cognitive abilities: proportion mediated=30.04%. The mediating role of punishment sensitivity was specific to the emotional/internalizing. This mediating role of both reward sensitivity and cognitive abilities was focusing on the behavioral/externalizing and neurodevelopmental dimensions of psychopathology.

**Conclusion:** We provide a better understanding of how genetic risks for MDD and ADHD confer risks for psychopathology and suggest potential prevention/intervention targets for children at-risk.

## Introduction

The last decade has seen major advances in psychiatric genetics. However fundamental questions remain about the psychological and cognitive links that associate genes with psychopathology^1^. Here, we examined whether three psychological and cognitive mechanisms-similar to those proposed by the National Institute of Mental Health’s Research Domain Criteria (RDoC)^2^ - punishment sensitivity, reward sensitivity and cognitive abilities - mediate the relationship between genetic liability and psychiatric symptoms.

Genetic liability to psychiatric disorders, e.g., major depressive disorder (MDD), attention-deficit/hyperactivity disorder (ADHD) and schizophrenia, can be captured by a composite of common gene variants identified from genome-wide association studies (GWAS), known as polygenic scores (PS)^3^. Multiple gene variants associated with different psychiatric disorders have been identified, and these cut across current diagnostic classification, such that genes contributing to one disorder also influence other phenotypes, not necessarily specific to such disorder^3–5^. For instance, PSs associated with case-status for MDD^4^, ADHD^3, 4^ and schizophrenia^4, 5^ in GWAS discovery studies are associated with covariation among multiple psychiatric symptoms not limited to their respective disorders, assessed through a latent variable known as the psychopathology factor or *p* factor^6^. It is unclear, however, what the mechanisms that explain this link between PSs and the *p* factor might be.

These mechanisms are likely to occur at multiple levels, including molecular^7^, cellular^8^, circuit^9^, but also psychological and cognitive^6^. The RDoC has emerged as a framework to investigate how components of these different levels contribute to psychopathology^2^. Surprisingly little work has been done to examine which psychological and cognitive mechanisms mediate links between genetic risks and psychopathology. This is unfortunate as 1) at least some psychological and cognitive phenotypes can be measured with relatively high precision^10–12^, and 2) some may also be amenable to interventions^13–15^. Understanding such mediating mechanisms may provide a foundation for effective early prevention and intervention strategies, an especially important goal in childhood given that most psychiatric disorders originate early in life. For this reason, our study focused on uncovering psychological and cognitive pathways between genetic liability (as indexed by PS), and psychopathology (as indexed by *p* factor as well as more specific latent variables, such as internalizing/emotional, externalizing/behavioral and neurodevelopmental) in children.

Here we study three psychological and cognitive mechanisms that show promise as potential mediators. The first two are related to motivation: punishment and reward sensitivity, or how easily affected individuals are by aversive and appetitive stimuli, respectively. Decades of work in psychopathology have demonstrated the importance of motivation^16^. Many emotional/internalizing symptoms (such as those that typify MDD, the depression-state in bipolar disorders and anxiety disorders) are associated with punishment sensitivity^17, 18^, as measured by the behavioral inhibition system (BIS) subscale^19^. Conversely, neurodevelopmental and behavioral/externalizing symptoms (such as those observed in ADHD and conduct disorders^20, 21^) are associated with reward sensitivity, as measured by the behavioral activation system (BAS) subscale^19^. High reward sensitivity, reflected by BAS, is also viewed as a major risk factor for bipolar disorders^22^. Accordingly, it is possible, for instance, that the previously found association between MDD PS and the *p* factor^4^ might be mediated through punishment sensitivity (the BIS) while the association between the ADHD PS and the *p* factor^3, 4^ might be mediated through reward sensitivity (the BAS).

The third mechanism is related to cognitive abilities. Cognitive abilities have many forms, ranging from executive functions^23^, verbal aptitude^24^, learning and memory^25^ to spatial reasoning^26^. The shared variance across different forms of cognitive abilities is represented by a latent variable, called the general intelligence factor, or *g* factor^27^. The *g* factor has been associated with multiple different dimensions of psychopathology^6, 11, 28^. A recent twin study^28^ also demonstrated that the *g* factor broadly captures genetic propensity for psychopathology. Accordingly, we hypothesized that the *g* factor may mediate between the genetic liability of various PSs and the *p* factor.

Recently the *p* factor has been integrated into a transdiagnostic framework that empirically groups related symptoms together in a hierarchical order of dimensions^29, 30^. This framework has advantages over classical diagnostic systems (e.g., *DSM*, or *International Statistical Classification of Diseases*, [ICD]) in terms of predicting clinical outcomes^31^, such as new onsets of future diagnoses^32^, suicide attempts^33^ and psychosocial impairments^34^. This hierarchical structure has the *p* factor at its apex to represent broad severity across different types of psychopathology^35^. Moreover, the *p* factor is manifested by lower, specific dimensions. Based on a recent large-scale study in children from the Adolescent Brain Cognitive Development (ABCD) dataset^30^, at the lower level of this hierarchical structure are five specific dimensions of behavioral/externalizing, neurodevelopmental, emotional/internalizing, somatoform and detachment items. These lower-level, specific dimensions make it possible to study mediating processes in greater granularity. Researchers can test which of the specific dimensions (in addition to the *p* factor) are mediated by each mechanism. For instance, while PS for one disorder and the *p* factor might be jointly mediated by two mechanisms, these two mechanisms might mediate different specific dimensions from the other, suggesting dissociable roles in the pathway from genetic risk to psychopathology.

Our aim here is to test possible missing mediating links between genetic risk and clinical symptoms. We do this by testing the hypotheses that motivational traits and cognitive abilities mediate between polygenic scores (PSs) for major psychiatric disorders and psychopathology (as indexed by the *p* factor and its underlying dimensions). The first step was to identify which of the common psychiatric PSs were associated with the *p* factor. Here we regressed the *p* factor on PSs for common psychiatric disorders: MDD, ADHD, anxiety, bipolar, schizophrenia and autism. We used PSs of specific disorders as opposed to cross disorders^36^ to demonstrate specific mediators for specific genetic risks for each disorder. Once identified, we conducted mediation analyses. While using mediation analyses on cross-sectional data does not provide causal mechanisms, mediation analyses still allow us to examine whether the variance of the relationship between each identified PS and the *p* factor is explained by the proposed mediators: punishment sensitivity (the BIS), reward sensitivity (the BAS) and cognitive abilities (the *g* factor). Finally, to further investigate the specific roles of the mediators, we conducted follow-up mediation analyses on the five specific dimensions by which the *p* factor was manifested. This allowed us to demonstrate detailed pathways for each mechanism to mediate specific sets of psychopathology.

## Method

### Sample

We used the baseline, cross-sectional data from ABCD Release 3, collected at 21 sites across the United States between September 1, 2016, and February 15, 2020 (http://dx.doi.org/10.15154/1519007). The study recruited 11,099 children across races and ethnicities^37^. Given the biases associated with generating PSs in samples that are ancestrally diverse, in line with others, we restricted our main analysis to children of homogeneous ancestry^38^. To match with the discovery samples of the PSs (assessed by multidimensional scaling analysis of their genotype data, see below), we focused our main analysis on children of European ancestry. After additional quality controls (see below), the final sample included 4814 children (2,263 female children; M_age_= 9.94 (SD=.61) years). We also separately conducted an exploratory, supplemental analysis on 1,460 children of African ancestry (726 female children; M_age_= 9.94 (SD=.60) years; see Figure S1, available online) using the same European ancestry-derived summary statistics. They were the second-largest population based on the genotype data in the dataset, but their ancestry did not match with the discovery samples of the PSs: these analyses were therefore exploratory, as recent work show lower predictive performance when non-matched ancestry samples are used^38^. The ABCD study was approved by the IRB at multiple sites and obtained informed consent (parents) and assent (child)^39^.

### Polygenic Score (PS)

Full details of genotyping have been published elsewhere^40^ (http://dx.doi.org/10.15154/1519007). Briefly, the study used the Smokescreen array and genotyped from saliva and whole blood. The ABCD applied quality control based on calling signals and variant call rates and performed the Ricopili pipeline. The study imputed the quality-controlled genotype data with TOPMED reference (https://topmedimpute.readthedocs.io/en/latest/prepare-your-data/). We excluded children whose genetic data were collected from a problematic plate or had a subject-matching issue based on the study’s recommendations. We further quality controlled using the genotypeqc function (https://github.com/ricanney/stata). We removed individuals with minimal or excessive heterozygosity, disproportionate levels of individual missingness (>2%) or insufficient sample replication (IBD < 0.8). We also excluded SNPs based on minor allele frequency (<5%), call rate (< 98%) or evidence for violations of Hardy-Weinberg equilibrium (*p* < 1E-10). We only included children with low genetic relatedness (more than 3rd-degree relative pairs; IBD ≥ 0.0422). We considered children to be genetically similar to the ancestry reference if they were within 4 standard deviations of the mean of the top 4 principal components (PCs) for the super-population individuals in the 1000 genomes Phase 3 reference genotypes (see Figure S2-S3, available online, for population structure PCs). After extracting children of the ancestry reference, we recomputed multidimensional scaling.

Using PLINK 1.9 via the summaryqc2score function (https://github.com/ricanney/stata), we computed each PS as the z-scored, weighted mean number of risk alleles in approximate linkage equilibrium, derived from imputed autosomal SNPs. We defined these risk alleles as those associated with case-status in large-scale discovery GWASs of six major psychiatric disorders: MDD^41^, ADHD^42^, anxiety^43^, bipolar^44^, schizophrenia^45^ and autism^46^. Note that for MDD, we used summary statistics based on diagnoses by clinicians^41^ and not based on self-reports used in 23andMe and UK Biobank as implemented in a more recent meta-analysis^47^. For anxiety, we used a GWAS that involved a meta-analysis from various populations^43^, as opposed to a more recent, larger study that only analyzed veterans^48^, which may not be generalizable to other populations.

In the main analysis, we focused on risk alleles that passed the *p* < .05 threshold in the discovery GWASs to capture most of the variance in moderately powered GWAS^49^. As an exploratory, supplementary analysis, we also used PSs at other thresholds from *p* < .5 to .0001 and applied Benjamini-Hochberg false discovery rate (FDR) to control for multiple testing across thresholds. In our structural equation modelling (SEM) that involved PSs, we also included control variables: (i) four principal components (to control for population stratification) and (ii) sex.

### Psychopathology: *p* factor and Five Specific Dimensions

We assessed children’s psychopathology using the Child Behavior Checklist (CBCL),^50^ reported by parents as detailed previously^51^. The CBCL included 119 items on a scale of 0 (not true) to 2 (very true or often true) that reflected emotional, behavioral and ADHD problems occurring in the past 6 months. Following previous work^30^, we removed low-frequency items and created composites for items that were highly correlated with each other. We captured the *p* factor and its lower, specific dimensions as latent variables in two confirmatory factor analysis (CFA) models.

First, the higher-order *p* factor model (Figure 1A) allowed us to model the *p* factor in the mediation analyses. Here we had the *p* factor as the 2^nd^-order latent variable and the five specific dimensions (behavioral/externalizing, neurodevelopmental, emotional/internalizing, somatoform and detachment; as defined previously^30^) as the 1^st^-order latent variables (i.e., the *p* factor was manifested by the five specific dimensions).

**Figure 1.**
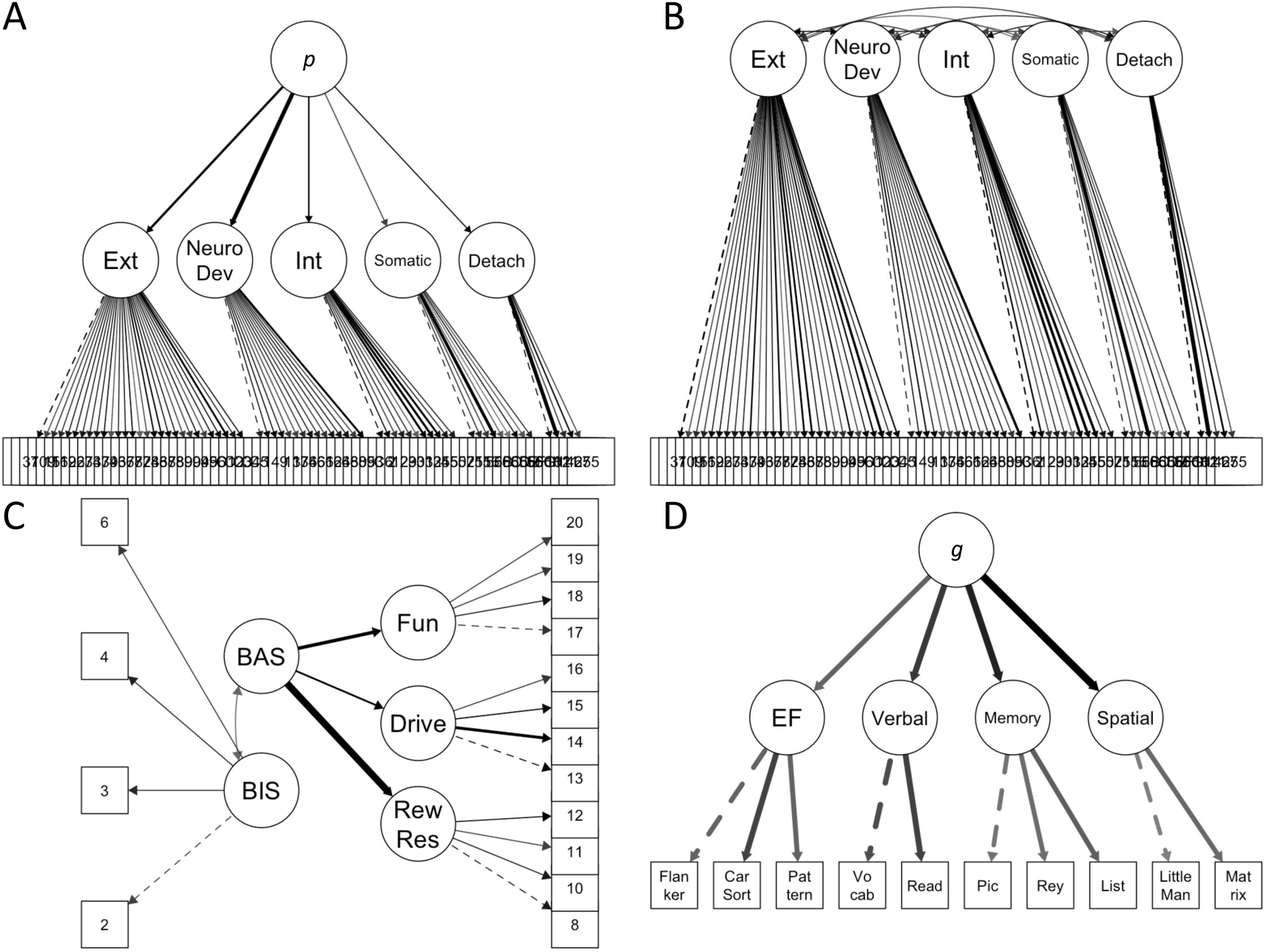
Confirmatory Factor Analysis for Latent Variables of Interest. **Note:** Line thickness reflects the magnitude of standardized parameter estimates. The dotted lines indicate marker variables that were fixed to 1. Numbers in squares indicate the item number from each scale. 1A and 1B show the higher-order *p* factor and first-order models for psychopathological phenotypes, respectively. The higher-order *p* factor showed the following fit indices: scaled CFI=.926, TLI=.924 and RMSE=.028 (90%CI=.027-.028). The *p* factor in this model also had good reliability: OmegaL2=.880. The first-order model also showed the following fit indices: scaled CFI=.930, TLI=.927 and RMSE=.027 (90%CI=.027-.028). Overall, its first-order variables had good reliability (total Omega3=.963) while reliability for specific dimensions varied: externalizing (Omega3=.927), neurodevelopmental (Omega3=.875), internalizing (Omega3=.840), somatoform (Omega3=.721) and detachment (Omega3=.655). 1C shows the BIS/BAS model. This model showed the following fit indices: robust, scaled CFI=.940, TLI=.928 and RMSE=.048 (90%CI=.045-.051). The latent variables had good reliability: 1^st^-order variables (total Omega3=.87) and the BAS (OmegaL2=.835). 1D shows the higher-order *g* factor model. This model showed the following fit indices: robust, scaled CFI=.965, TLI=.949 and RMSE=.046 (90%CI=.041-.050). The *g* factor had good reliability: OmegaL2=.865. BAS = behavioral approach system; BIS = behavioral inhibition system; Car Sort = card sort; Detach = detachment; Drive = BAS drive; EF = executive functions; Ext = externalizing; Fun = BAS fun; *g* = *g* factor; Int = internalizing; List = list sorting working memory; Matrix = matrix reasoning; Neuro Dev = neurodevelopmental; *p* = *p* factor; Pattern = pattern comparison processing; Pic = picture sequence memory; Read = oral reading recognition; Rew Res = BAS reward responsiveness; Rey = Rey-auditory verbal learning; Somatic = somatoform; Vocab = picture vocabulary.

Second, the first-order model (Figure 1B) allowed us to model the five specific dimensions as correlated latent variables in the follow-up mediation analyses. We had the five specific dimensions as the 1^st^-order, correlated latent variables without the *p* factor. Using this model, we could test associations between variables (PSs and mediators) and each of the five specific dimensions while controlling for the correlations among the specific dimensions.

### Motivation: Punishment (BIS) and Reward (BAS) Sensitivity

We assessed children’s motivation through the behavioral inhibition system and behavioral activation system (BIS/BAS) scale^19^ modified from PhenX, reported by children as detailed previously^51^. The scale included 20 items (7 for BIS) on 4-point Likert options (0=not true; 3=very true) that reflected punishment (BIS) and reward (BAS) sensitivity. The scale has been developed to have a 4-factor structure: one BIS and three BAS subscales (fun, drive, reward responsiveness)^19^. Following a recent factor analysis in children^52^, we dropped four problematic items: three from the BIS and one from the BAS-reward-responsiveness. This resulted in four items per (sub)scale. To evaluate the latent structure of the BIS/BAS, we ran a CFA model, similar to the classical^19^ and revised^52^ 4-factor models. In this model (Figure 1C), BAS (as the higher-order variable) underlined the three BAS subscales (as the 1^st^-order variables). We then allowed the BIS and BAS to covary. We treated the BIS and BAS as latent mediators for the mediation analyses.

### Cognitive Abilities: *g* Factor

We assessed children’s cognitive abilities through various cognitive tasks as detailed previously^11^. Children completed these tasks on an iPad during a 70-min in-session visit. We included 10 tasks in our model (including seven tasks from the NIH Toolbox^12^). First, the Flanker task measured inhibitory control. Second, the card sort task measured cognitive flexibility. Third, the pattern comparison processing task measured processing speed. Fourth, the picture vocabulary task measured language and vocabulary comprehension. Fifth, the oral reading recognition task measured language decoding and reading. Sixth, the picture sequence memory task measured episodic memory. Seventh, the Rey-auditory verbal learning task measured auditory learning, recall and recognition. Eight, the list sorting working memory task measured working-memory^12^. Ninth, the little man task measured visuospatial processing via mental rotation^26^. Tenth, the matrix reasoning task measured visuospatial problem solving and inductive reasoning^25^.

As a preliminary analysis, we followed previous work^11^ by applying principal component analysis (PCA) to evaluate the structure of cognitive abilities. As we assume some similarity among the tasks, we used oblique (oblimin) rotations. The 4-component solution appeared to capture the cognitive tasks well, given minimal cross-loading. From this preliminary analysis, we then used CFA to capture the latent variable, the *g* factor, the underlying cognitive abilities. In our higher-order *g* factor model (Figure 1D), we had the *g* factor as the 2^nd^-order latent variable. We also had four 1^st^- order latent variables in the model: executive functions (capturing the Flanker, card sort and pattern comparison processing tasks), verbal (capturing the picture vocabulary and oral reading recognition tasks), memory (capturing the picture sequence memory, Rey-auditory verbal learning and list sorting working memory tasks) and spatial (capturing the little man and matrix reasoning tasks). We treated the *g* factor as a latent mediator for the mediation analyses.

### Statistical Approach: Mediation Analyses with Structural Equation Modeling

In our mediation analyses, we fit a series of latent variable models in successive steps. First, to identify which of the PSs are associated with the *p* factor, we treated the *p* factor from the higher-order *p* factor model as an outcome variable and the six PSs (MDD, ADHD, anxiety, bipolar, schizophrenia and autism) as explanatory variables. In this regression SEM model, the association between each PS and *p* factor was already controlled for other PSs. Second, to ensure that the proposed mediators were related to the *p* factor, we treated the *p* factor as an outcome variable and the three proposed mechanisms (the BIS, the BAS and the *g* factor) as explanatory variables. Third, to demonstrate which of the three mediators were related to the PSs implicated by the first step, we treated the three proposed mediators as outcome variables and each of the selected PSs as an explanatory variable. Fourth, we examined the extent to which the relationship between each of the selected PS and the *p* factor was accounted for by the mediators implicated by steps two and three. Here we treated each implicated PS (step one) as an independent variable, mechanisms (steps two and three) as mediators and the *p* factor as a dependent variable. We then conducted follow-up analyses to further examine the role of the mediators by exploring associations with the five specific dimensions from the first-order model. For these follow-up mediation analyses, we started by examining the association between the five specific dimensions (as outcome variables) and each of the PS that was significantly associated with the *p* factor (as an explanatory variable). Because the first-order model separately estimated correlations among the five specific dimensions, here we captured the unique associations between each specific dimension and PS (i.e., controlling for the correlations among the dimensions). Only the five specific dimensions of psychopathology that were associated with each PS were used in the final follow-up mediation analyses. Finally, we tested the indirect effects, or how much the relationship between each significant PS and the specific dimensions of psychopathology was accounted for by the mediators. Given that we used specific dimensions as multiple endogenous (i.e., dependent) latent variables, we further controlled for multiple-testing by applying FDR to all joint indirect effects that included all mediators for each specific dimension. For latent variable modeling configurations, see Supplement 1, available online. For the R script for data pre-processing and latent variable modeling and their detailed outputs, see https://narunpat.github.io/MotivationCognitionMediationPolygenicScores/MovCogMedPSPFactor_ABCD3_TestWGender_PC4_Mac.html.

## Results

### How Well Do the Proposed Latent Variable Models Fit the Data?

Figure 1 shows the results of confirmatory factor analysis (CFA). All of the four proposed latent variable models had adequate model fit indices. Overall, the proposed latent variables (including the *p* factor, five specific dimensions, BIS, BAS, *g* factor) had good reliability (i.e., internal consistency), reflected by OmegaL2^53^ for 2^nd^-order variables and Omega3^54^ for 1^st^-order variables.

### Which PSs are Associated with the *p* Factor?

Our first SEM tested the relationship between the six psychiatric PSs and the *p* factor from the higher-order *p* factor model on children of European ancestry (Figure 2A, 2B). At *p*<.05 PS threshold, only the MDD and ADHD PSs showed unique associations with the *p* factor. When examining the associations at different PS thresholds, we found that the *p* factor was significantly associated with the ADHD PS across all six PS thresholds, but was only significantly associated with the MDD PS at four PS thresholds (*p*<.5 to <.01). Given (1) that the MDD and ADHD PSs showed associations with the *p* factor at a similar magnitude at the pre-specified *p*<.05 PS threshold and (2) that these associations passed the FDR correction, we treated the MDD and ADHD PSs as independent variables in our subsequent mediation analyses on children of European ancestry. Note that we also conducted the same SEM on children of African ancestry at *p*<.05 PS threshold (see S3, available online). However, none of the six psychiatric PSs were significantly associated with the *p* factor in this population. Accordingly, we did not conduct further mediation analyses on children of African ancestry.

**Figure 2.**
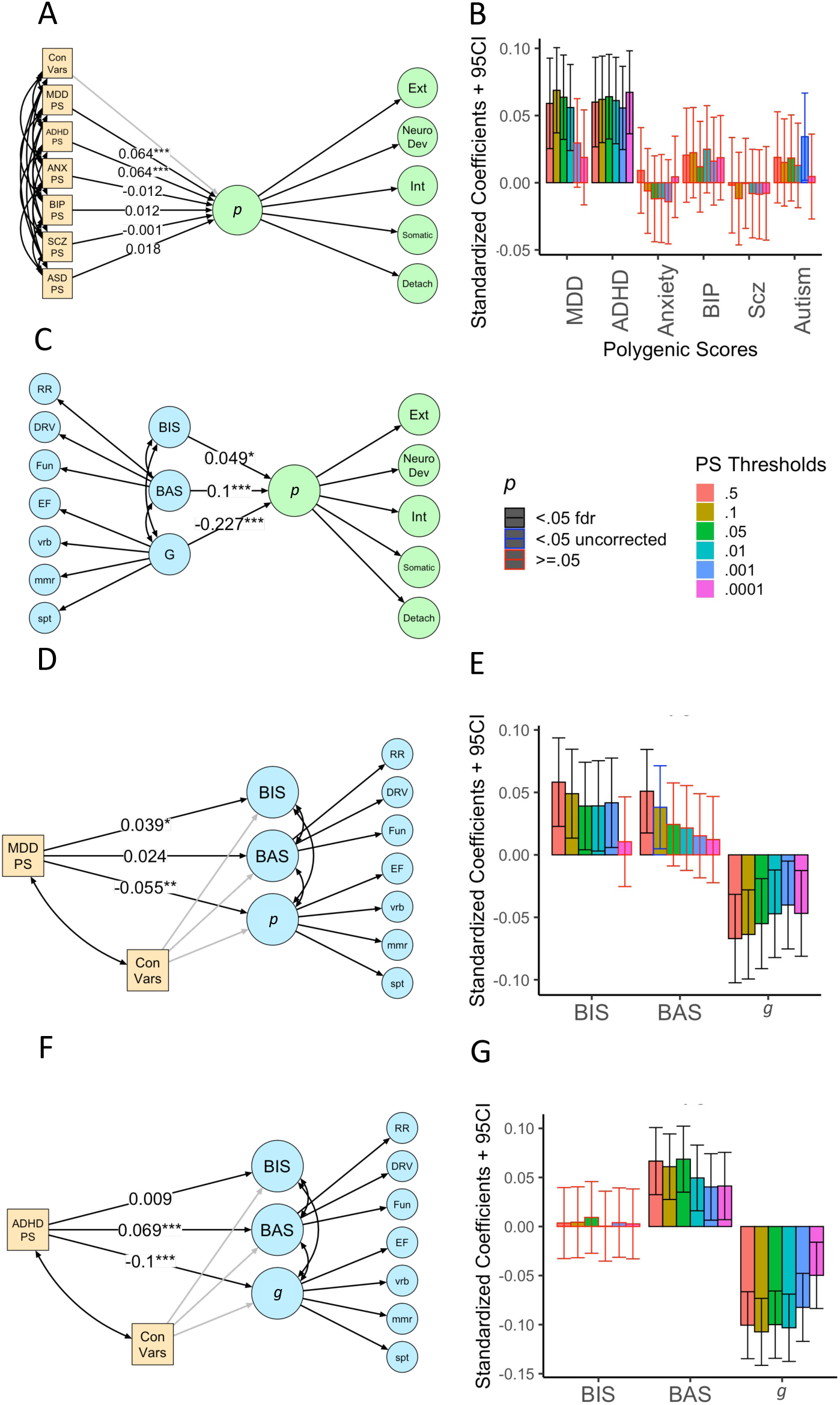
The Relationships Between Polygenic Scores (PS), the *p* factor and the Proposed Mediators (Behavioral Inhibition System [BIS], Behavioral Activation System [BAS] and *g* Factor) **Note:** A shows SEM testing the relationship between PSs and the *p* factor. 2A shows SEM testing the relationship between six PSs and the *p* factor using p<.05 PS threshold. This model demonstrated the following fit indices: robust, scaled CFI=.940, TLI=.938 and RMSE=.032 (90%CI=.031-.032). From this model, the MDD (β=.064, SE=.016, 95%CI = 0.032 - 0.095, z=3.972, *p*<.001) and ADHD (β=.064, SE=.016, 95%CI = 0.033 - 0.096, z=3.983, *p*<.001) PSs, but not other PSs, showed unique associations with the *p factor*. These relationships survived FDR correction when examining PSs across thresholds (2B). 2C shows SEM testing the relationship between the proposed mediators and the *p* factor. This model demonstrated the following fit indices: scaled CFI=.920, TLI=.918 and RMSE=.022 (90%CI=.022 - .023). The *p* factor was significantly associated with all proposed mediators: the BIS (β=.049, SE=.025, 95%CI = 0005 - .098, z=1.979, *p*=.048), BAS (β=.1, SE=.024, z=4.144, 95%CI = .053 - .147, *p*<.001) and *g* factor (β=-.227, SE=.021, 95%CI = -.269 - -.185, z=-10.571, *p*<.001). 2D shows SEM testing the relationship between MDD PS and the proposed mediators using p<.05 PS threshold. This model demonstrated the following fit indices: robust, scaled CFI=.952, TLI=.946 and RMSE=.032 (90%CI=.030-.033). From this model, MDD PS was significantly related to the BIS (β=.039, SE=.018, 95%CI = .004 - .074, z=2.186, *p*=.026) and the *g* factor (β=-.055, SE=.018, 95%CI = -.091 - .019, z=-2.996, *p*=.003), but not the BAS (β=.024, SE=.017, 95%CI = -.009 - .058, z=1.436, *p*=.151). These relationships survived FDR correction when examining PSs across thresholds (2E). 2F shows SEM testing the relationship between the ADHD PS and the proposed mediators using *p*<.05 PS threshold. This model demonstrated the following fit indices: robust, scaled CFI=.952, TLI=.946 and RMSE=.032 (90%CI=.030-.033). From this model, ADHD PS was significantly related to the BAS (β=.069, SE=.017, 95%CI = .035 - .102, z= 4, *p*<.001) and the *g* factor (β=-.1, SE=.018, 95%CI =-.134 - -.066, z=-5.714, *p*<.001), but not the BIS (β=.009, SE=.019, 95%CI = -.027 - .046, z=.495, *p*=.621). These relationships survived FDR correction when examining PSs across thresholds (2G). The numbers indicate standardized parameter estimates. Yellow indicates proposed independent variables. Green indicates dependent variables. Blue indicates proposed mediators. ADHD = attention deficit hyperactivity disorder; ASD = autism spectrum disorder; BAS = behavioral approach system; BIP = bipolar disorder; BIS = behavioral inhibition system; Con Vars = PS control variables (four principal components and sex); Detach = detachment; DRV = BAS drive; EF = executive functions; Ext = externalizing; FDR = false discovery rate; Fun = BAS fun; G = *g* factor; Int = internalizing; MDD = major depressive disorder; mmr = memory; Neuro Dev = neurodevelopmental; PS = polygenic score; RR = BAS reward responsiveness; SCZ = schizophrenia; Somatic = somatoform; spt = spatial; vrb = verbal. \**p*<.05; \*\**p*<.01; \*\*\**p*<.001.

### Are the Proposed Mechanisms Related to the *p* Factor?

Next, we evaluated whether the proposed mediators were related to the main dependent variable, *p* factor (Figure 2C). Here, we examined the *p* factor in relationship to the BIS, BAS and *g* factor simultaneously, again allowing for the assessment of unique relationships. The *p* factor was significantly associated with all proposed mediators: the BIS, BAS and *g* factor.

### Are the Proposed Mechanisms Related to MDD and ADHD PSs?

We then separately evaluated whether each of the PSs that were associated with the *p* factor (MDD and ADHD PSs) were also related to each of the proposed mediators at *p*<.05 PS threshold. MDD PS (Figure 2D, 2E) was significantly related to the BIS and the *g* factor, but not the BAS. The BIS and the *g* factor were therefore included as mediators for MDD PS mediation analyses. ADHD PS (Figure 2F, 2G) was significantly related to the BAS and the *g* factor, but not the BIS. The BAS and the *g* factor were therefore included as mediators for the ADHD PS mediation analyses.

### Do the Proposed Mechanisms Mediate between Each PS and the *p* Factor?

We then conducted the mediation SEM separately for MDD and ADHD PSs at *p*<.05 PS threshold. For the MDD PS mediation model, the BIS and *g* factor were included as mediators (Figure 3A, 3B). The association between MDD PS and the *p* factor was partially mediated by both the BIS (proportion mediated=5.73%) and the *g* factor (proportion mediated=16.60%), together explaining 22.35% of the association.

**Figure 3.**
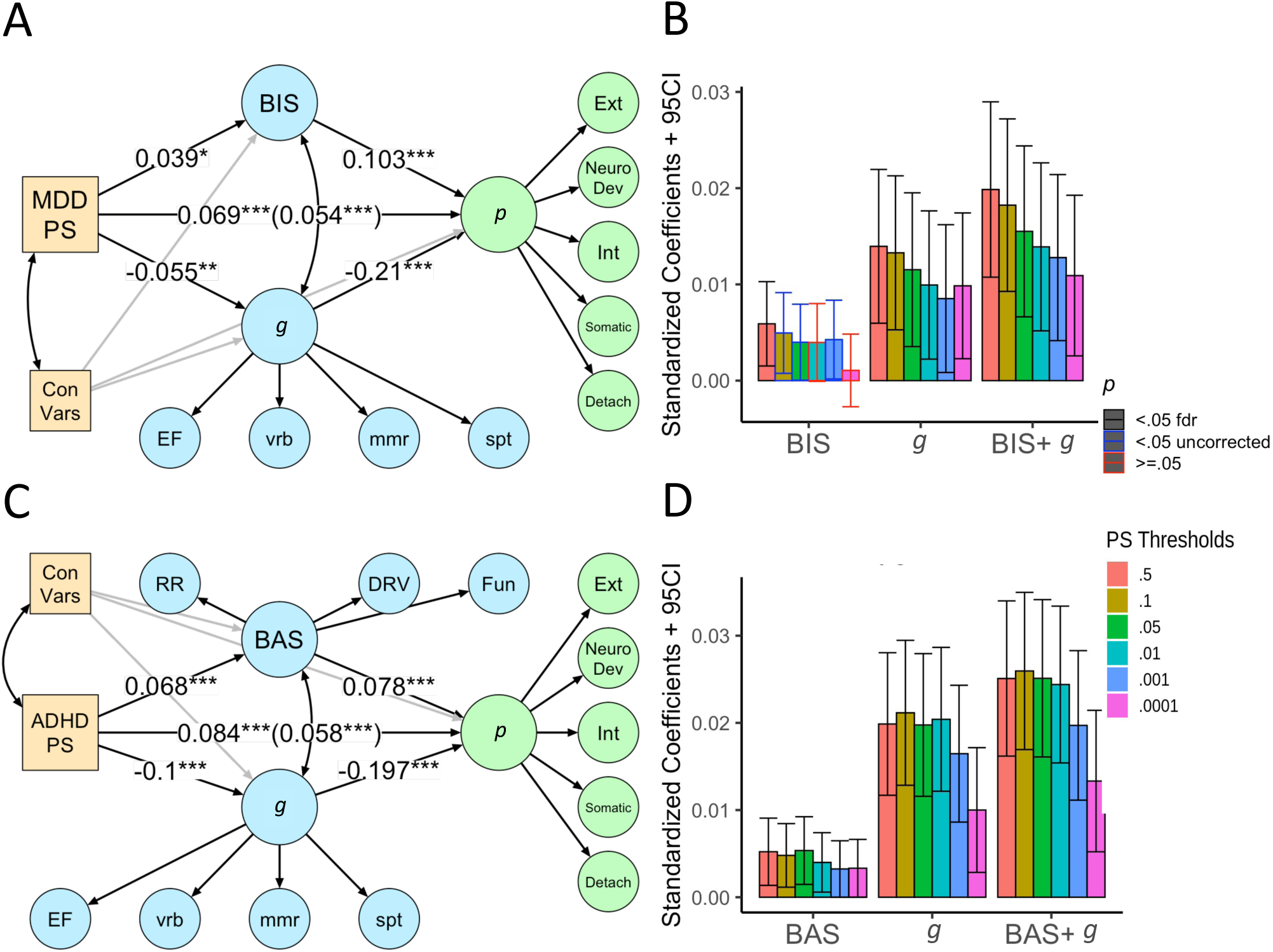
The Mediations Between Polygenic Scores (PS) (Major Depressive Disorder [MDD] and Attention-Deficit/Hyperactivity Disorder [ADHD]) and the *p* Factor. **Note:** 3A shows structural equation modelling (SEM) testing the mediation between MDD PS and the *p* factor with the BIS and *g* factor as mediators using *p*<.05 PS threshold. This model demonstrated the following fit indices: robust, scaled CFI=.81, TLI=.804 and RMSE=.034 (90%CI=.033-.034). From this model, the association between MDD PS and the *p* factor was partially mediated by both the BIS (indirect β=.004, SE=.002, 95CI% = .00003 - .008, z=1.976, *p*=.048) and the *g* factor (indirect β=.012, SE=.004, 95CI% = .004 - .02, z=2.826, *p*=.005), making the joint indirect effect at β=.015 (SE=.005, 95CI% = .007 - .024, z=3.424, *p*=.001). The *g* factor and joint indirect effect at *p*<.05 PS threshold survived FDR correction when examining PSs across thresholds (3B). While the BIS indirect effect was significant (*p*<.05) at multiple PS thresholds (.5, .1, .05, .001), only the effect at *p*<.5 PS threshold survived FDR correction. 3C shows SEM testing the mediation between the ADHD PS and the *p* factor with BAS and *g* factor as mediators using *p*<.05 PS threshold. This model demonstrated the following fit indices: robust, scaled CFI=.825, TLI=.821 and RMSE=.032 (90%CI=.031-.032). From this model, the association between the ADHD PS and *p* factor was partially mediated by both the BAS (indirect β=.005, SE=.002, 95%CI = .001 - .009, z=2.704, *p*=.007) and *g* factor (indirect β=.020, SE=.004, 95%CI = .012 - .028, z=4.731, *p*<.001), making the joint indirect effect at β=.03 (SE=.005, 95CI% = .016 - .034, z=5.451, *p*<.001). The BAS, *g* factor and joint indirect effect at *p*<.05 PS threshold survived FDR correction when examining PSs across thresholds (3D). The numbers indicate standardized parameter estimates. The parenthesis indicates the direct effect between each of the PS and the *p* factor after accounted for by the mediators. Yellow indicates independent variables. Blue indicates mediators. Green indicates dependent variables. ADHD = attention deficit hyperactivity disorder; BAS = behavioral approach system; BIS = behavioral inhibition system; Con Vars = PS control variables (four principal components and sex). Detach = detachment; DRV = BAS drive; EF = executive functions; Ext = externalizing; FDR = false discovery rate; Fun = BAS fun; G = *g* factor; Int = internalizing; MDD = major depressive disorder; mmr = memory; Neuro Dev = neurodevelopmental; PS = polygenic score; RR = BAS reward responsiveness; Scz = schizophrenia; Somatic = somatoform; spt = spatial; vrb = verbal. \**p*<.05; \*\**p*<.01; \*\*\**p*<.001.

The ADHD PS mediation model included the BAS and *g* factor as mediators at *p*<.05 PS threshold (Figure 3B). The association between the ADHD PS and *p* factor was partially mediated by both the BAS (proportion mediated=6.404%) and *g* factor (proportion mediated=23.637%). Thus, the two mediators together explained 30.040% of the association between the ADHD PS and *p* factor.

### Which of the Five Specific Dimensions are Associated with Each PS?

We then conducted follow-up mediation analyses to investigate the distinct roles of the mediators at the level of five specific dimensions for both the MDD and ADHD PSs. We first tested the relationship between each of the two PSs and the five dimensions at *p*<.05 PS threshold. The MDD PS was significantly associated with all five specific dimensions (see Figure 4A, 4B): all dimensions were therefore included in the follow-up mediation analyses for MDD PS.

**Figure 4.**
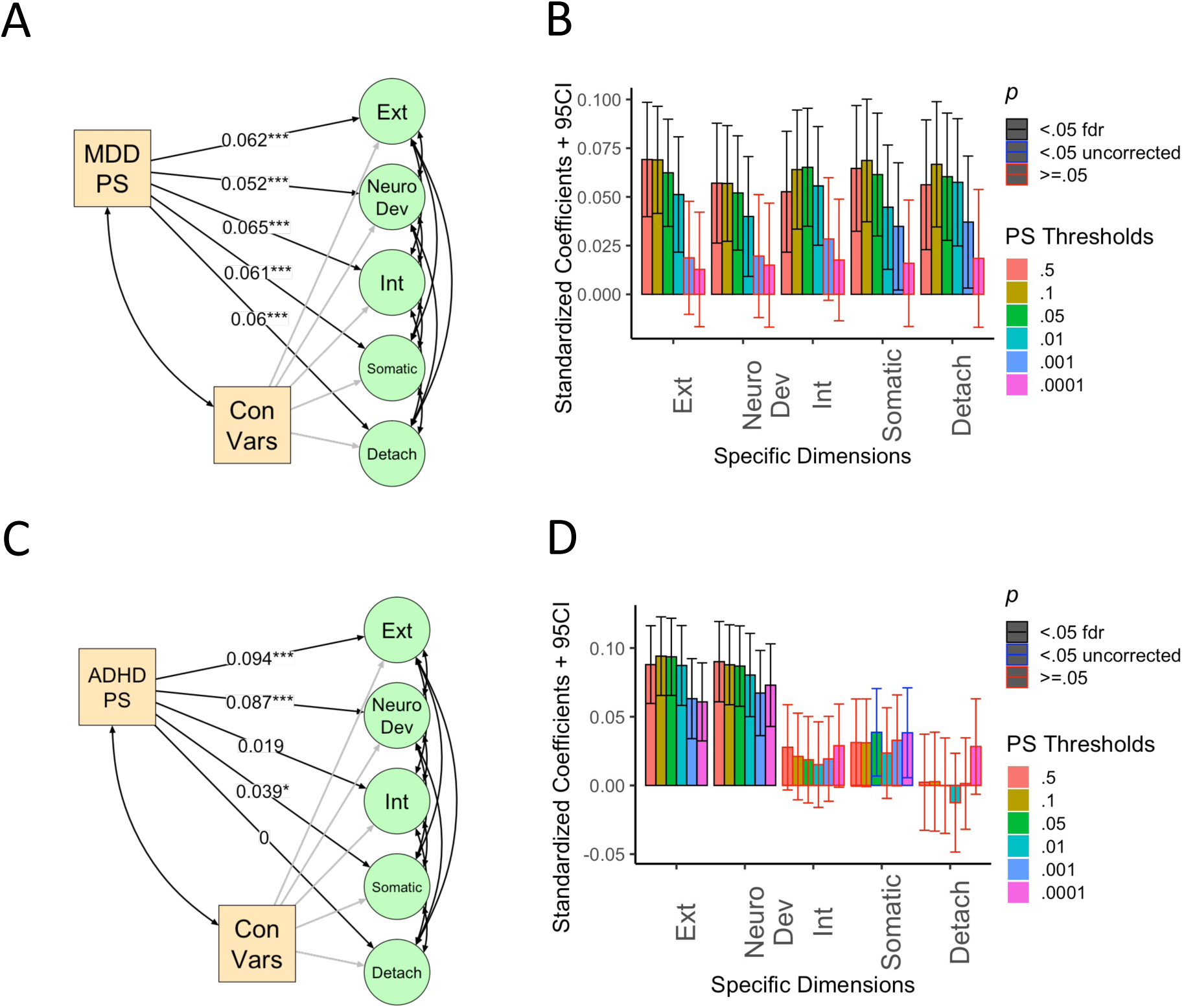
The Relationships Between Polygenic Scores (PS) (Major Depressive Disorder [MDD] and Attention-Deficit/Hyperactivity Disorder [ADHD]) and Specific Dimensions of Psychopathology. Note: 4A shows SEM testing the relationship between MDD PS and the five specific dimensions from the first-order model using *p*<.05 PS threshold. This model demonstrated the following fit indices: robust, scaled CFI=.79, TLI=.781 and RMSE=.038 (90%CI=.037-.039). From this model, MDD PS was significantly related to all five specific dimensions. These relationships survived FDR correction when examining PSs across thresholds (4B). 4C shows SEM testing the relationship between the ADHD PS and the five specific dimensions from the first-order model using *p*<.05 PS threshold. This model demonstrated the following fit indices: robust, scaled CFI=.79, TLI=.781and RMSE=.038 (90%CI=.037-.039). From this model, ADHD PS was significantly related to externalizing, neurodevelopmental and somatoform but not internalizing and detachment. The relationships with externalizing and neurodevelopmental (but not somatoform) survived FDR correction when examining PSs across thresholds (4D). The numbers indicate standardized parameter estimates. Yellow indicates independent variables. Green indicates dependent variables. ADHD = attention deficit hyperactivity disorder; Con Vars = PS control variables (four principal components and sex); Detach = detachment; Ext = externalizing; FDR = false discovery rate; *g* = *g* factor; Int = internalizing; MDD = major depressive disorder; Neuro Dev = neurodevelopmental; PS = polygenic score; Scz = schizophrenia; Somatic = somatoform. \**p*<.05; \*\**p*<.01; \*\*\**p*<.001.

The ADHD PS was statistically associated with externalizing, neurodevelopmental and somatoform but not internalizing and detachment (see Figure 4C, 4D): these three dimensions were therefore included in the follow-up mediation analyses for the ADHD PS.

### Do the Proposed Mechanisms Mediate between Each PS and Specific Dimensions?

For the MDD PS follow-up mediation model at *p*<.05 PS threshold (Figure 5A, 5B), joint indirect effects from all five specific dimensions passed the FDR correction (*p*_fdr_=.004-.018). The BIS specifically mediated the influence of MDD PS on internalizing (proportion mediated=13.715%). The *g* factor largely mediated the influence of MDD PS on externalizing (proportion mediated=18.082%) and neurodevelopmental (proportion mediated=32.237%) dimensions, but also on internalizing (proportion mediated=5.747%) and somatoform (proportion mediated=5.647%).

**Figure 5.**
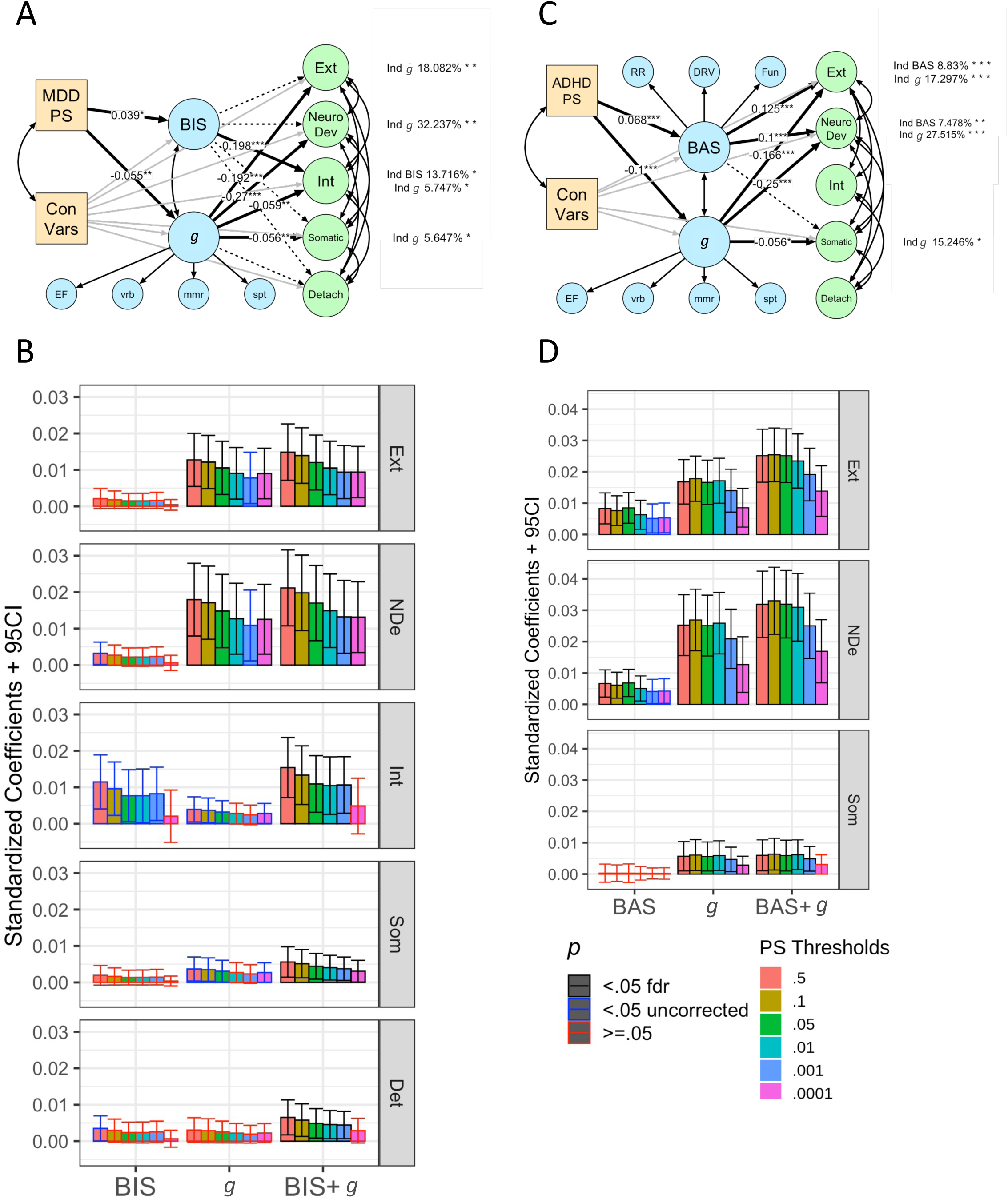
The Mediation Between Polygenic Scores (PS) (Major Depressive Disorder [MDD] and Attention-Deficit/Hyperactivity Disorder [ADHD]) and Specific Dimensions of Psychopathology. **Note:** 5A shows SEM testing the mediation between MDD PS and the five specific dimensions of psychopathology with the BIS and *g* factor as mediators using *p*<.05 PS threshold. This model demonstrated the following fit indices: robust, scaled CFI=.815, TLI=.807 and RMSE=.033 (90%CI=.033-.034). The BIS mediated the relationship between MDD PS and internalizing (indirect β=.008, SE=.004, 95%CI = .001 - .015, z= 2.112, *p*=.035). The *g* factor mediated the relationship between MDD PS and externalizing (indirect β=.011, SE=.004, 95%CI = .003 - .018, z= 2.831, *p*=.005), neurodevelopmental (indirect β=.015, SE=.0052, 95%CI = .005 - .025, z= 2.887, *p*=.004), internalizing (indirect β=.003, SE=.002, 95%CI = .000 - .006, z= 2.028, *p*=.043) and somatoform (indirect β=.003, SE=.002, 95%CI = .000 - .006, z= 2.026, *p*=.043). Only the indirect effects of the *g* factor on externalizing and neurodevelopmental survived the FDR correction when examining PSs across thresholds (5B). While BIS’s indirect effect on internalizing was significant (*p*<.05) at multiple PS thresholds (.5, .1, .05, .01, .001), none survived the FDR correction. 5C shows SEM testing the mediation between ADHD PS and the three specific dimensions of psychopathology with BAS and *g* factor as mediators using p<.05 PS threshold. This model demonstrated the following fit indices: robust, scaled CFI=.809, TLI=.803 and RMSE=.033 (90%CI=.033-.034). The BAS mediated the influence of the ADHD PS on externalizing (indirect β=.008, SE=.002, 95%CI = .004 - .013, z= 3.399, *p*=.001) and neurodevelopmental (indirect β=.007, SE=.002, 95%CI = .002 - .011, z= 3.078, *p*=.002). The *g* factor mediated the influence of the ADHD PS on all three dimensions: externalizing (indirect β=.017, SE=.004, 95%CI = .010 - .024, z= 4.597, *p*<.001), neurodevelopmental (indirect β=.025, SE=.005, 95%CI = .015 - .035, z= 5.072, *p*<.001) and somatoform (indirect β=.006, SE=.002, 95%CI = .001 - .010, z= 2.381, *p*=.017). All of these indirect effects survived the FDR correction when examining PSs across thresholds (5D). The numbers overlaid black lines indicate standardized parameter estimates. The numbers on the right side next to each specific dimension of psychopathology indicate proportion mediated for the mediation paths with significant indirect effects. Dotted lines indicate mediation paths with non-significant (*p*<.05) indirect effects. Yellow indicates independent variables. Blue indicates mediators. Green indicates dependent variables. ADHD = attention deficit hyperactivity disorder; BAS = behavioral approach system; BIS = behavioral inhibition system; Con Vars = PS control variables (four principal components and sex); Detach = detachment; DRV = BAS drive; EF = executive functions; Ext = externalizing; FDR = false discovery rate; Fun = BAS fun; *g* = *g* factor; Int = internalizing; MDD = major depressive disorder; mmr = memory; Neuro Dev = neurodevelopmental; PS = polygenic score; RR = BAS reward responsiveness; Scz = schizophrenia; Somatic = somatoform; spt = spatial; vrb = verbal. \**p*<.05; \*\**p*<.01; \*\*\**p*<.001.

For the ADHD PS follow-up mediation model at *p*<.05 PS threshold (Figure 5C, 5D), joint indirect effects from the three included specific dimensions passed the FDR correction (*p*_fdr_= <.001-.019). The BAS mediated the influence of the ADHD PS on externalizing (proportion mediated=8.83%) and neurodevelopmental (proportion mediated=7.478%), whereas the *g* factor mediated the influence of the ADHD PS on all three dimensions: externalizing (proportion mediated=17.297%), neurodevelopmental (proportion mediated=27.515%) and somatoform (proportion mediated=15.246%).

## Discussion

Here we aimed to uncover the psychological and cognitive mechanisms mediating the relationship between genetics and psychopathology. In particular, we tested whether three RDoC- based psychological and cognitive mechanisms, namely punishment sensitivity (BIS), reward sensitivity (BAS) and cognitive abilities (*g* factor), mediated the relationship between polygenic scores (PSs) for different psychiatric disorders and psychiatric symptoms across disorders (the *p* factor and its specific dimensions). We first identified that, among the six common psychiatric PSs, MDD and ADHD PSs were associated with the *p* factor in children. While we did not find a previously shown relationship between schizophrenia PS and the *p* factor^4, 5^, MDD and ADHD PSs were associated with the *p* factor, consistent with the previous reports^3, 4^. Moreover, MDD and ADHD PSs were related to our proposed mediation mechanisms. Importantly, the proposed mechanisms partially mediated the relationship of the two PSs to the *p* factor and its specific dimensions. Note that our observation that no associations were observed in children of African ancestry is in keeping with other studies that have observed low predictive power of PSs derived from a discovery sample when the target sample is of different ancestry^38^.

The relationship of MDD PS to the *p* factor was mediated by the BIS and *g* factor, whereas the relationship of ADHD PS to the *p* factor was mediated by the BAS and *g* factor. Thus, the influence of MDD and ADHD PSs on psychopathology may be acting through both shared (*g* factor) and unique (BIS versus BAS) routes. Here we demonstrated two routes for MDD PS (punishment sensitivity and cognitive abilities) and two routes for ADHD PS that are partially dissociable from MDD PS (reward sensitivity and cognitive abilities). To further investigate the specificity of these pathways, we conducted follow-up mediation analyses on the five specific dimensions of psychopathology by which the *p* factor was manifested. As discussed in more detail below, our results showed that the proposed psychological and cognitive mechanisms differentially mediated each of the five specific dimensions. Thus, together these data are consistent with the hypothesis that dissociable but complementary pathways mediate the influence of the MDD and ADHD PSs on the *p* factor.

The mediating role of the BIS from MDD PS to the *p* factor is consistent with studies associating the BIS with emotional/internalizing symptoms^17^. When examining its detailed mediating pathways using specific dimensions of psychopathology, we found a high level of specificity in the mediation: the BIS was only significantly related to one PS (MDD PS) and one specific dimension of psychopathology (internalizing). Conversely, the mediating role of the BAS to ADHD PS and the *p* factor is consistent with associating the BAS with neurodevelopmental and behavioral/externalizing symptoms^20, 21^. Similar to the BIS, the BAS also showed a high level of specificity in its mediation: it was only significantly related to one PS (ADHD PS) and two specific dimensions (neurodevelopmental and behavioral/externalizing symptoms). Together, these findings suggest that motivation-related mechanisms, punishment and reward sensitivity, mediated the influences of genetics in a specific manner.

Of note, we found relatively broad influences of the MDD PS on all specific dimensions of psychopathology, including both internalizing and externalizing dimensions. Yet, the MDD PS was specifically related to punishment sensitivity (BIS), but not reward sensitivity (BAS). This seems to suggest that other mediators may play a role in the relationship between the MDD PS and other psychopathological dimensions beyond internalizing. The *g* factor appears to be one of these mediators. The mediating role of the *g* factor to both the MDD and ADHD PSs is in line with previous work showing relationships between cognitive abilities and broad psychopathology^6, 11^. Unlike the two motivation-related mediators, the *g* factor showed a broader role. That is, the *g* factor mediated the influences between 1) both the MDD and ADHD PSs and 2) various specific dimensions of psychopathology. For both the MDD and ADHD PS, the *g* factor strongly mediated the contribution of genetic influences to the externalizing and neurodevelopmental dimensions, relative to other dimensions. The *g* factor additionally, albeit weakly, mediated the link with the internalizing and somatic dimensions for the MDD PS and with the somatic dimension for the ADHD PS. Accordingly, we found that having genetic liability for MDD and/or ADHD had negative associations with cognitive abilities, which in turn, may enhance the general risk to develop psychopathology. As such, cognitive abilities played a key role as a non-specific factor that linked genetic liability with broad psychopathology, consistent with previous findings and theoretical perspectives of the *p* factor^6, 35^.

We believe understanding the roles of the proposed psychological and cognitive mechanisms has research and clinical implications. As we showed here, the model fit indices and reliability (i.e., internal consistency) indices of the proposed mechanisms were relatively high. This means that we can measure these mechanisms in children with precision using latent variable modeling. Moreover, punishment and reward sensitivity and cognitive abilities are shown to be altered via psychotherapy and other environment-altering interventions^13–15^. Accordingly, they can be targeted for effective early prevention and intervention strategies for children at risk.

This study is not without limitations. First, as highlighted previously^55^, while using PSs derived from GWAS are more reliable than using only a few common SNPs from selected genes (the candidate-gene approach), PSs still explain only a small proportion of genetic liability to psychiatric disorders. The relatively small effect sizes of our results confirm this notion. Additionally, our use of CBCL^50^ to define psychopathology did not allow us to investigate psychosis as another specific dimension. Thus, our definition of the *p* factor may not be exhaustive. This may explain the non-significant relationship between schizophrenia PS and the *p* factor, which is contradictory to previous studies^4, 5^. Next, we measured the mediators and psychopathology at the same time, making it difficult to empirically test the directionality of their relationships. Fortunately, the ABCD is an ongoing longitudinal study that will provide additional data from the same children until they are 20 years old. Thus, we believe that our study will lay a foundation for future research to further empirically test the directionality of the effects found here, for instance, using the cross-lagged panel model. Further, we only included children with low genetic relatedness (more than 3rd-degree relative pairs) to avoid inflated associations following recommendations^38, 56^. Nonetheless, this method may lower the statistical power. Future studies may implement a different approach to statistically account for relatedness without exclusion.

Finally, the generalizability of the findings is limited to children of European ancestry due to the lack of summary statistics from well-powered GWASs for major psychiatric disorders done in non-European participants^57^. When we applied European ancestry-derived summary statistics to children with African ancestry, we no longer saw the relationship between psychiatric PSs and the *p* factor. This is consistent with recent work showing lower predictive performance when non-matched ancestry samples are used^38^. This highlights the importance of having diverse populations in the GWASs and statistical approaches to deal with multi-ancestry and admixed cohorts^57^, so that genetic research can be more boardy applicable. Such shortcomings unfortunately prohibited the full utilisation of the ABCD study even though the ABCD study had been specifically designed to have a strength in the diversity of its participants^37^.

In summary, in a large sample of children, the influences of genetic predispositions for MDD and ADHD on psychopathology were mediated by three RDoC mechanisms: punishment sensitivity, reward sensitivity and cognitive abilities. These findings further our understanding of the structure of psychopathology and the pathways through which it relates to genetic architecture.

## Data Availability

Data used in the preparation of this article were publically available. The data can be obtained from the Adolescent Brain Cognitive Development (ABCD) Study (https://abcdstudy.org), held in the NIMH Data Archive (NDA).

## Acknowledgments

Data used in the preparation of this article were obtained from the Adolescent Brain Cognitive Development (ABCD) Study (https://abcdstudy.org), held in the National Institute of Mental Health (NIMH) Data Archive (NDA). This is a multisite, longitudinal study designed to recruit more than 10,000 children age 9-10 and follow them over 10 years into early adulthood. The ABCD Study is supported by the National Institutes of Health (NIH) and additional federal partners under award numbers U01DA041022, U01DA041028, U01DA041048, U01DA041089, U01DA041106, U01DA041117, U01DA041120, U01DA041134, U01DA041148, U01DA041156, U01DA041174, U24DA041123, U24DA041147, U01DA041093, and U01DA041025. A full list of supporters is available at https://abcdstudy.org/federal-partners.html. A listing of participating sites and a complete listing of the study investigators can be found at https://abcdstudy.org/scientists/workgroups/. ABCD consortium investigators designed and implemented the study and/or provided data but did not necessarily participate in analysis or writing of this report. This manuscript reflects the views of the authors and may not reflect the opinions or views of the NIH or ABCD consortium investigators. NP, YW and AS were supported by Otago Medical Research Foundation Grant though M Begg Charitable Trust.

## Supplement 1

### Latent Variable Modelling Configurations

For each CFA structure, we fixed latent factor variances to one, so that we could estimate all factor loadings. We used robust estimators to deal with the non-normality of psychopathological phenotypes in this population-based study. To this end, we first used robust maximum likelihood estimation (MLR) with robust (Huber-White) standard errors and scaled test statistics that also dealt with missing values via the Full Information Maximum Likelihood algorithm. However, if we encountered a non-convergent problem with the MLR, we treated data as ordinal and used WLSMV as an estimator instead. The WLSMV uses diagonally weighted least squares to estimate model parameters. To demonstrate model fit, we used scaled comparative fit index (CFI), Tucker-Lewis Index (TLI) and root mean squared error of approximation (RMSEA) with 90% CI. We reported the robust versions of these indicators for the MLR. For CFA, we also reported the reliability of the latent variables: OmegaL2^1^ for 2^nd^- order variables and Omega3^2^ for 1^st^-order variables. These reliability indices reflect the internal consistency of the latent variables of interest. Model fits and reliability indices are shown in the figure captions. We ran the analyses in R4.0.2 on the standardized data using lavaan^3^ (version=.6-6) and semTools^1^ along with semPlot^4^ and qgraph^5^ for visualization (see https://narunpat.github.io/MotivationCognitionMediationPolygenicScores/MovCogMedPSPFact or_ABCD3_TestWGender_PC4_Mac.html for the script and detailed outputs).

**Figure S1.**
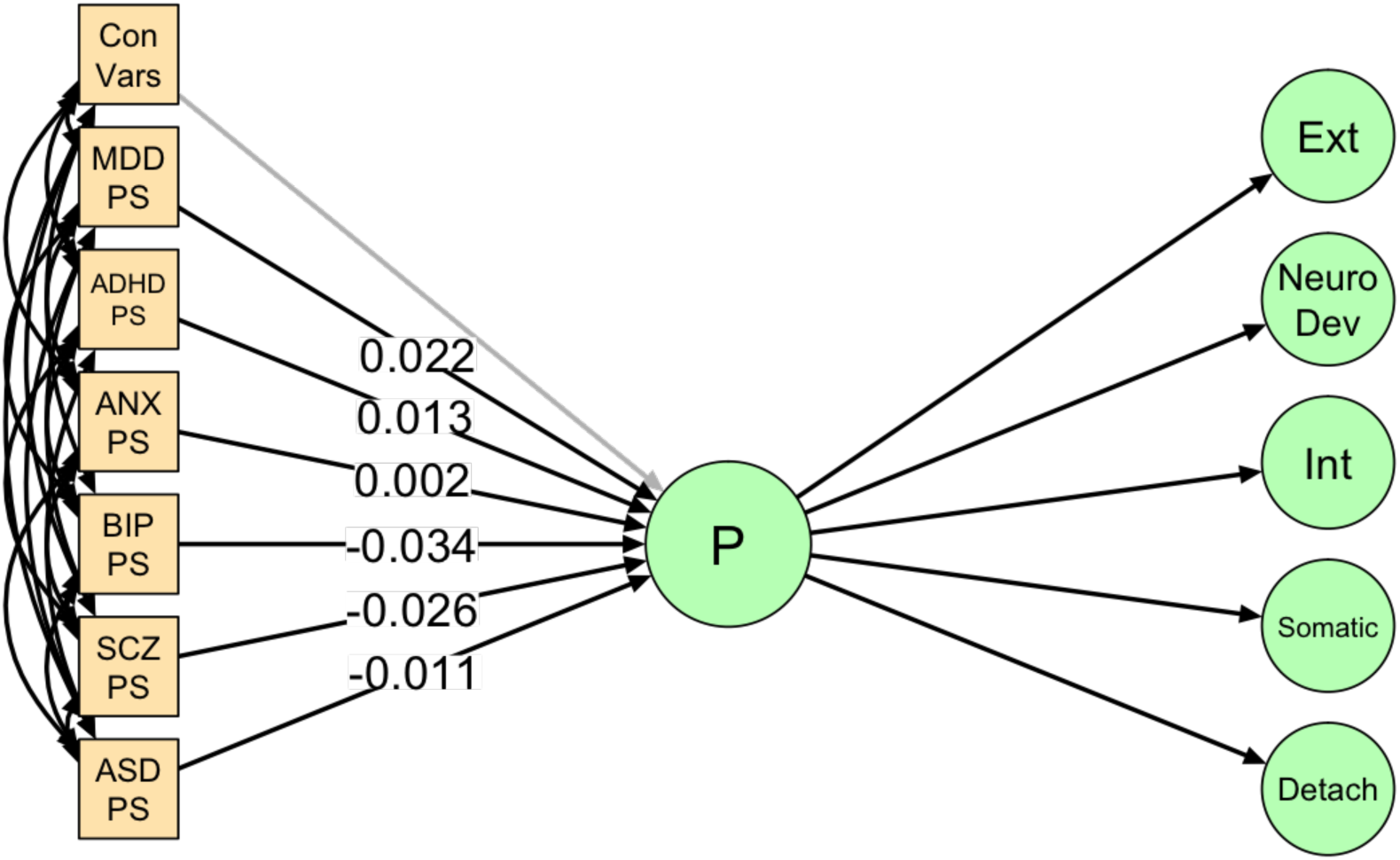
The relationships between polygenic scores (PSs) and the *p* factor among Children of African ancestry. This structural equation model (SEM) model using *p*<.05 PS threshold showed the following fit indices: robust, scaled CFI=.795, TLI=.790 and RMSE=.038 (90%CI=.037- .039). From this model, none of the six PSs showed significant associations with the *p* factor at *p* < .05. ADHD = attention deficit hyperactivity disorder; ASD = autism spectrum disorder; BIP = bipolar disorder; Con Vars = PS control variables (four principal components and sex); Detach = detachment; Ext = externalizing; Int = internalizing; MDD = major depressive disorder; Neuro Dev = neurodevelopmental; P = *p* factor; PS = polygenic score; SCZ = schizophrenia; Somatic = somatoform; *p<.05; **p<.01; ***p<.001.

**Figure S2.**
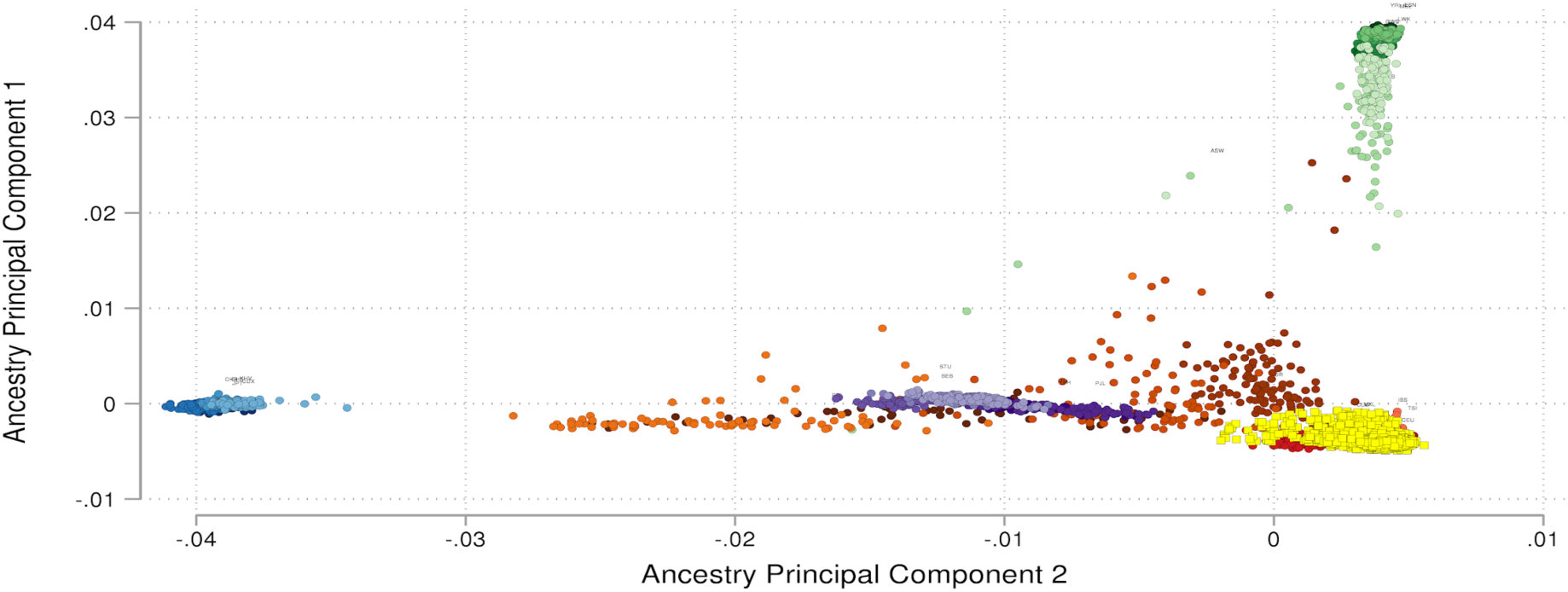
Population Structure Principal Components for Children of European Ancestry in the Adolescent Brain Cognitive Development (ABCD) dataset. We used the super-population individuals in the 1000 genomes Phase 3 as reference genotypes. Red cluster = European super- population; Orange = Americas super-population; Green = Africa super-population; Blue = East Asia super-population; Purple = South Asia super-population; Yellow = Children of European Ancestry in the ABCD.

**Figure S3.**
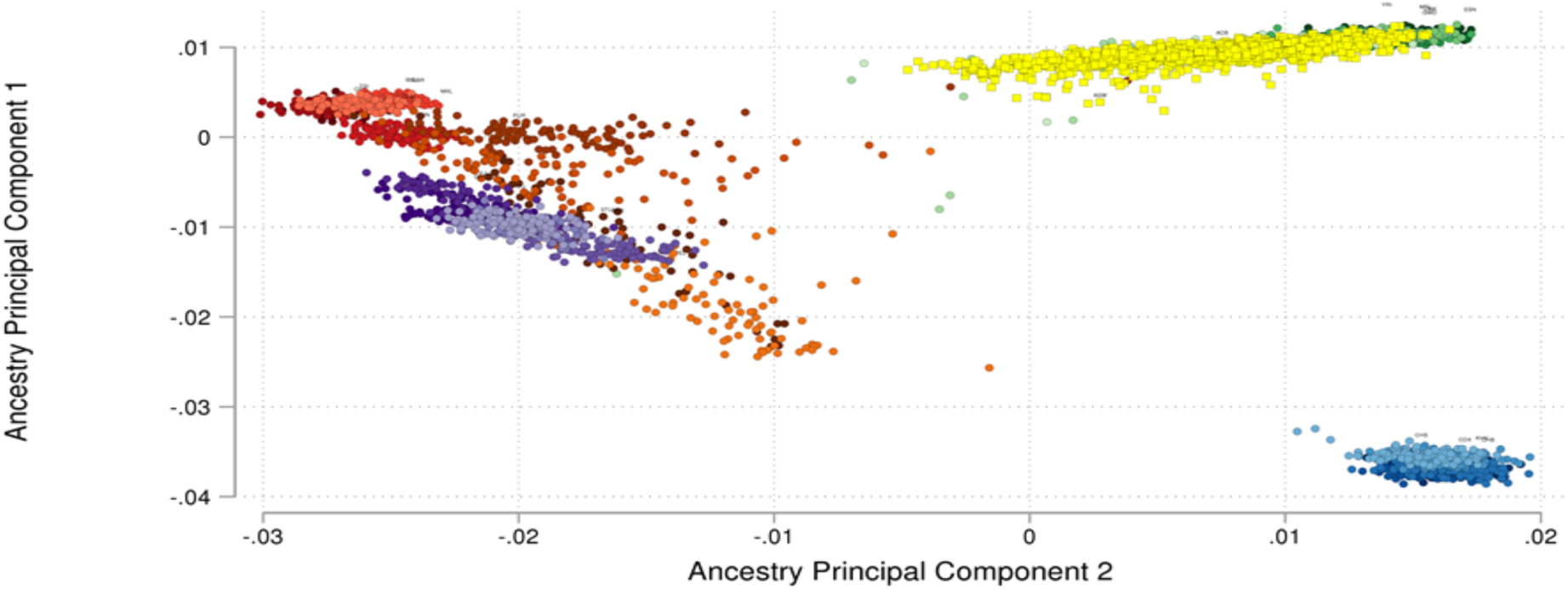
Population Structure Principal Components for Children of African Ancestry in the Adolescent Brain Cognitive Development (ABCD) dataset. We used the super-population individuals in the 1000 genomes Phase 3 as reference genotypes. Red cluster = European super-population; Orange = Americas super-population; Green = Africa super-population; Blue = East Asia super-population; Purple = South Asia super-population Yellow = Children of African ancestry in the ABCD.

